# Functional and Organic Pseudo-Orthostatic Tremor: Clinical Features, Electrophysiology, and Treatment Outcomes in a Retrospective Cohort

**DOI:** 10.64898/2026.10.18.26365840

**Authors:** Agustín Querejeta-Coma, Luis Yupanqui-Guerra, Laura Santos-Sánchez-de-las-Matas, Adriana Gómez-Domínguez, Héctor Canfield-Medina, Roberto Cano-de-la-Cuerda, Carlos Ordás-Bandera

## Abstract

**Background:** Pseudo-orthostatic tremor (psOT) is a rare form of tremor of the lower body, triggered by standing, at frequencies lower than 13 Hz. It is believed to be secondary to other neurological disorders in a large proportion of cases. Surface electromyography (sEMG) can identify its core characteristics and differentiate it from other similar disorders such as primary orthostatic tremor (POT) and orthostatic myoclonus (OM).

**Objectives:** Our study aims to illustrate the clinical and neurophysiological features as well as the outcomes and final diagnoses of patients with psOT in a retrospective two-centre cohort.

**Methods:** We retrospectively examined the available clinical and neurophysiological records of 35 consecutive patients with a diagnosis of psOT in the 2022-2026 period. Data on demographics, clinical characteristics, sEMG findings, pharmacological treatments, ancillary investigations and patient reported outcomes were collected.

**Results:** 35 patients with psOT were identified, of which 6 (17%) displayed features consistent with a functional neurological disorder (FND). Among the 29 patients with organic psOT, 21 (72.4%) fulfilled the diagnostic criteria for essential tremor (ET), while an identifiable neurological cause was established in four (13.8%). sEMG findings differed between functional and organic psOT, with the latter often displaying features consistent with weight-bearing hyperkinesias.

**Conclusions:** Our retrospective study suggests that psOT is most often associated with ET, while an identifiable neurological cause was found in a relatively small proportion of patients. We also describe functional psOT as a previously unrecognized clinical presentation of FND.

## Introduction

The term orthostatic tremor (OT) was first coined in 1984 to designate a tremor of the lower body that is triggered by standing at high frequencies, ranging from 13 to 18 Hertz (Hz) [1]. Prior accounts of this phenomenon date to Pazzaglia‚s work in 1970 [2]. OT is typically alleviated by walking or leaning and interrupted by seating or lying down [3]. In the late 1980s, tremors displaying similar phenomenology at frequencies <10 Hz were described in some patients with essential tremor (ET), and were initially termed slow orthostatic tremors [4] [5]. Other neurophysiological features, such as coherence between muscle pairs, tend to be dissimilar between fast and slow tremors, hence the term “pseudo-orthostatic tremor” (psOT) has been reserved since the 2018 consensus for any tremors whose frequencies are lower than 13 Hz [6].

Most often, classical OT is an isolated tremor syndrome that comes unaccompanied by any other neurological features, hence referred to as primary orthostatic tremor (POT). When it happens in combination to other neurological condition it is labelled as primary orthostatic tremor plus (POT-plus) [6]. These views have been challenged by recent works that have identified that substantial proportions of patients with OT actually suffer from combined tremor syndromes [7] and/or relevant neurological comorbidities [8], [9]. In contrast, most psOT are seen as secondary [10] to several other non-neurological [11] and neurological disorders, such as degenerative parkinsonisms [12], [13] or hereditary cerebellar ataxias [14]. Some authors have even suggested that psOT may not exist as a primary entity [15], but this has been the matter of controversy [10]. psOT are, however, regarded as rare phenomena and have been the subject of a limited number of publications in the last years.

The diagnosis should be secured by surface electromyography (sEMG) which will allow precise identification of the phenomena [10], that may be difficult (if often not impossible) to achieve with the standard neurological examination [16]. sEMG can confidently demonstrate the presence of a tremor, investigate its coherence, burst duration and contraction patterns as well as its response to weight transfer from the lower limbs, which would confirm a *weight-bearing* mechanism when the tremor alleviates, often being transmitted to the upper limbs [3]. Also, it allows the identification of orthostatic myoclonus (OM), a common mimic of OT and psOT [17]. sEMG may as well have the potential to identify features of internal inconsistency that are typical of functional neurological disorders (FND) although this has not been studied yet in patients that display tremor during standing.

To date, a number of relatively small retrospective series have included patients with psOT. In 2019 a review of the available literature on the topic was published by Hassan et al. [10] comprising 70 cases of electrophysiologically confirmed ‘slow OT‚: 33% were isolated tremor syndromes, 67% had a coexistent disorder that could explain the tremor and only 2% had a formal diagnosis of ET. The median frequency range was 6-7 Hz and coherence and synchronicity of the bursts, which are characteristic features of fast OT, varied significantly amongst the reports. Since, further retrospective case series have been reported [9], [18] widening the understanding of psOT. In particular, the study by Bicart-See [9] identified cerebellar and/or extrapyramidal signs in all of the 17 psOT patients included, with Parkinson‚s Disease (PD) as the most frequent cause, in 53% of the patients.

Our study aims to illustrate the clinical and neurophysiological features as well as the outcomes and final diagnoses of a two-centre case retrospective cohort of psOT that includes, for the first time, the description of patients with psOT secondary to FND.

## Methodology

### Design

This retrospective observational study was designed and reported in accordance with the Strengthening the Reporting of Observational Studies in Epidemiology (STROBE) guidelines [19].

The research was approved by the Research Ethics Committee of the Fundación Jiménez Díaz University Hospital (EO243-25_HRJC) and was conducted in accordance with the principles of the Declaration of Helsinki and applicable national regulations. Given the retrospective nature of the study and the use of previously collected clinical data, the requirement for informed consent was waived by the Ethics Committee. All data were anonymized prior to analysis to ensure participant confidentiality and privacy.

All study data were collected and managed using REDCap electronic data capture tools [20].

### Patients

We retrospectively examined the clinical and neurophysiological records of 35 consecutive patients with a diagnosis of psOT, defined as tremor of the lower limbs at frequencies lower than 13 Hz that was present during standing, evaluated between January 2022 and August 2026 at the Rey Juan Carlos Hospital (Madrid, Spain) and Infanta Elena Hospital (Madrid, Spain). Patients with demonstrable resting tremor of the lower limbs were excluded.

Patients were classified as having functional or organic psOT according to their final clinical diagnoses. The diagnosis of functional psOT was established during the initial neurophysiological evaluation by a movement disorders neurologist, based on positive signs of internal inconsistency demonstrated during sEMG, and confirmed during clinical follow-up. Final diagnostic classification incorporated the clinical and neurophysiological findings available throughout follow-up.

### Baseline demographics

Data on patient gender, age of onset, diagnostic delay and total time of follow up were collected.

### Prior medical and medication history

We gathered information on pre-existing diagnoses of movement disorders, gait disorders or any other relevant neurological and non-neurological comorbidities. In addition, we searched for family history of tremor and/or PD in first or second degree relatives.

Medications charts were reviewed in the look for continued use of medications, in a compatible timeframe, that could cause or exacerbate tremor, as judged by the investigators.

### Symptoms and examination findings

The presence of subjective complaints prior to the diagnosis was retrieved from the clinical records and recorded in a pre-specified systematic manner for all patients focusing on lower limb tremor, trunk tremor, unsteadiness, lower limb subjective weakness, falls, fear of falling without falls, back pain, orthostatic intolerance as well as head, voice and face tremors.

A similar approach was used for the presence of examination findings. The investigators systematically searched the clinical records looking for observations of lower limb tremor, Romberg‚s sign, unsteady gait, parkinsonism, upper limb action and/or resting tremor, myoclonus, head and/or voice tremor, lower limb weakness and positive signs of FND.

Other symptoms or signs, if not pre-especified, could be registered in free text fields, when present.

### Neurophysiological features

For all patients we performed sEMG recordings using a 4-channel Natus device and surface electrodes in the four limbs. All patients had been advised to withdraw for 24 hours any medications prescribed to improved their tremors. Every patient at our centres is examined jointly by a neurophysiologist and a movement disorders specialist.

We reviewed the available reports for each patient and systematically recorded the dominant tremor frequency in Hz, the distribution of the tremor, its response to weight transfer (attenuation with weight transfer from the lower limbs and transmission to the upper limbs) as well as the presence of distractibility. Our group asseses distractibility looking at the response of the EMG trace during mental tasks (e.g. substracting 7s) and manual tasks (finger-to-finger test). A tremor is considered distractible if such tasks lead to a loss of the tremor structure.

If other, not previously specified, findings were encountered they could be registered in free text fields.

### Ancillary testing

Records were searched to retrieve the results of: 1) structural neuroimaging of the brain, either computerized tomography (CT) or magnetic resonance imaging (MRI); 2) DaTSCAN Single-Photon Emission Computed Tomography (DaTSCAN SPECT), 3) fluorodeoxyglucose positron emission tomography (FDG-PET) and 4) nerve conduction studies with needle electromyography (NCS/EMG)

### Final clinical diagnoses

We searched the final diagnoses amongst the most recent follow-up data. The available data were used to classify the patient‚s tremor syndrome (Axis I) and, when identifyable, its underlying etiology (Axis II) within the framework of the 2018 Consensus Statement on the classification of tremors [6]. The fulfillment of diagnostic criteria for essential tremor (ET) was assessed individually, independently of the presence of standing-induced lower-limb tremor.

Additionally, we gathered information on relevant neurological and non-neurological comorbidities, which were listed in free text fields.

### Outcomes

Patient reported outcomes were retrospectively rated using the Patient Global Impression of Change (PGI-C) using the most recent available data from follow-up. Current and previous use of antitremor agents was recorded. For descriptive purposes, discontinued medications were treated as ineffective although reasons for discontinuation were not systematically assessed. Consequently, this classification should not be interpreted as definitive evidence of treatment inefficacy. Medications still being taken at follow-up were classified as effective when the patient reported any degree of improvement on the PGI- C.

### Statistical analyses

Statistical analysis was performed on Jamovi Version 2.6.45.0 [21] and R-studio version 4.5.2 [22]. Data have been expressed as percentages (%) for categorical data and mean ± standard deviation (SD) or median and interquartile range (IQR) for Gaussian and non-Gaussian continuous data, respectively. Between-group comparisons, while considered exploratory due to the small simple size, were carried out with Fisher‚s exact test for categorical data, Student‚s t for Gaussian continuous data and Wilcoxon-Mann-Whitney for non-Gaussian.

Given the exploratory nature of the analyses, we did not correct for multiple comparisons.

**Results**

### Baseline demographics

Our search identified 35 consecutive patients with a diagnosis of psOT, 6 of which (17%) were classified as functional. Baseline demographics are detailed in table 1. Most patients were female (74%). The mean age of onset of psOT was 61 (SD =16). Median diagnostic delay was 40 months (IQR=72). Patients were followed up after diagnosis for 13 months on average (SD=11). There were no significant statistical differences between both groups.

**Table 1.** Baseline demographics of organic and functional pseudo-orthostatic tremor patients.

| Patients (n) | Organic pseudo-OT (29) | Functional pseudo-OT (6) | All (35) | *p*-value |
| --- | --- | --- | --- | --- |
| Female, n (%) | 22 (76) | 4 (67) | 26 (74) | 0.635 |
| Age of onset (years), mean ±SD | 63 ± 13 | 54 ± 28 | 61 ± 16 | 0.288 |
| Diagnostic delay (months), median [IQR] | 47 [70] | 29 [43] | 40 [72] | 0.429 |
| Follow-up time after diagnosis (months), mean ± SD | 11±10 | 20±13 | 13 ± 11 | 0.058 |

IQR: Interquartile range; OT: Orthostatic tremor; SD: Standard deviation

### Prior medical and medication history

15 patients in the organic psOT group (51.7%) had a pre-existing diagnosis of a movement disorder while none in the functional group did (p=0.027): 13 held a diagnosis of ET, 1 hemifacial spasm and 1 ET and FND tremor overlay. 4 patients (11%) had been diagnosed with gait disorders, 2 in each group (p=0.064): in the organic group, one patient had sensory ataxia attributed to a polyneuropathy and another had been diagnosed with “multifactorial unsteadiness”, whereas both patients in the functional group had gait disturbances attributed to chronic back pain syndromes, one associated to failed-back syndrome after numerous surgeries and the other secondary to a traffic accident.

A family history of tremor was reported in 15/19 (78.9%) organic cases with available information, corresponding to 15/29 (51.7%) of the entire organic group. A family history of PD was reported in 3/9 (33.3%) organic cases with available information, corresponding to 3/29 (10.3%) of the entire organic group. Family-history data were unavailable for all six functional psOT and for 10/29 and 20/29 organic cases for tremor and PD, respectively, precluding reliable direct comparisons between groups.

Other relevant neurological diagnoses included giant-cell arteritis, “orthostatic intolerance” and “MRI-negative myelopathy” in 3 functional patients (one each) and occipital neuralgia and transient ischaemic attack due to symptomatic carotid stenosis in 2 (one each) organic psOT patients.

Continued use of medications that could cause or exacerbate tremor before the onset of symptoms was documented in 18 patients (53%), 14 in the organic (50%) and 4 in the functional psOT group (67%) without any significant differences between the groups (p=0.47). No differences were found when comparing the use of beta-agonists (p=1), dopamine receptor blockers (p=1) and antidepressants between the groups (p=0.166). Supplementary Figure 1 illustrates prior medication usage across both groups. Supplementary table 2 provides a detailed account of exposure to individual medications.

### Symptoms and examination findings

Reported symptoms and observed signs prior to diagnosis are detailed in Supplementary Table 1.

Three subjective complaints were present in most subjects: lower limb tremor, unsteadiness and upper limb tremor. Whereas there were no significant differences in the frequency of reported subjective lower limb tremor (27 subjects, 77%) and unsteadiness (20 subjects, 57%), upper limb tremor was significantly more common in the organic psOT group (83 vs 33%, p=0.027). Interestingly, a tremor of the trunk was reported in 3 (50%) functional patients and none in the organic group (p=0.003).

In most subjects (30, 86%) a lower limb tremor could be identified in neurological examination. The presence of upper limb action tremor was also a common feature, present in 76% of organic and 50% of functional patients (p=0.322). Remarkably, positive signs of FND were reported in 4 (14%) and 3 (50%) of organic and functional psOT (p=0.079). In all of the former distractibility of lower limb tremor was suspected in clinical examination, although ultimately not confirmed with sEMG (see next section). In the functional psOT group one patient displayed a distractible hand tremor, another a combination of functional hemifacial spasm and head tremor and the remainder, a complex gait pattern that included antieconomic postures, leg dragging and knee buckling.

### Neurophysiological features

A lower limb tremor was present in all patients at standing with a median frequency of 6 Hz (IQR=2.5, range 4-10Hz) with no differences between groups (p=0.43). The tremor was bilateral in all but one patient while it was asymmetrically distributed in a minority in both groups (26%, p=1). 90% of organic psOT attenuated when weight was transferred from the lower limbs while this feature was absent (and often not recorded) in functional psOT patients (p = 0.022), all of which displayed disappearance and/or marked disruption of the tremor structure during distraction tasks.

Distractibility was documented during sEMG in all six patients with a final diagnosis of functional pseudo-orthostatic tremor, compared with none of the 18 patients with organic pseudo-orthostatic tremor for whom this information was available. As neurophysiological signs of internal inconsistency contributed to the diagnostic classification, this finding should be interpreted as a descriptive characteristic of the diagnostic groups rather than an independent measure of diagnostic discrimination.

### sEMG findings are summarised in table 2

The majority of organic psOT involved the calf/shin muscles whereas most of functional psOT involved the thigh compartment although there were no significant between-group differences in the individual muscles or muscle groups. Tremor distribution is detailed in Supplementary Figure 2.

**Table 2.** Surface electromyography findings in organic and functional pseudo-orthostatic tremor. Categorical data are presented as n/N (%), where N represents the number of patients with available data for each variable; percentages were calculated using available observations as the denominator. Hz: Hertz, OT: orthostatic tremor, sEMG: surface electromyography

| Patients (n) | Organic pseudo-OT (29) | Functional pseudo-OT (6) | All (35) | Missing (n) | *p*-value |
| --- | --- | --- | --- | --- | --- |
| Tremor (Hz), median, [IRQ] | 6 [2] | 7.5 [2.9] | 6 [2.5] | 0 | 0.428 |
| Bilateral tremor, n/N (%) | 28/29 (97) | 6/6 (100) | 34/35 (97) | 0 | 1 |
| Asymmetrical tremor, n/N (%) | 8/29 (28) | 1/6 (17) | 9/35 (26) | 0 | 1 |
| Response to weight transfer, n/N (%) | 26/29 (90) | 0/2 | 26/31 (84) | 4 | 0.022 |
| Transmission to the upper limbs, n/N (%) | 7/18 (39) | 0/3 | 7/21 (33) | 14 | 0.521 |
| Distractibility, n/N (%) | 0/18 (0) | 6/6 (100) | 6/24 (25) | 11 | <0.001 |

Other relevant findings included incongruent features in the functional psOT patients, namely delayed abolition of the lower limb tremor after sitting in 1 patient, tremor increase with weight transfer in 2 patients. Within the organic psOT group: 1 patient displayed concomitant OM that appeared right after standing before EMG bursts were organised in a rhythmical manner (i.e. tremor) and, in 2 patients, there were features suggestive of functional tremor in the upper limbs but not in the lower limbs, therefore interpreted as possible functional overlay.

Representative examples of sEMG recordings are provided in Figure 1.

**Figure 1.**
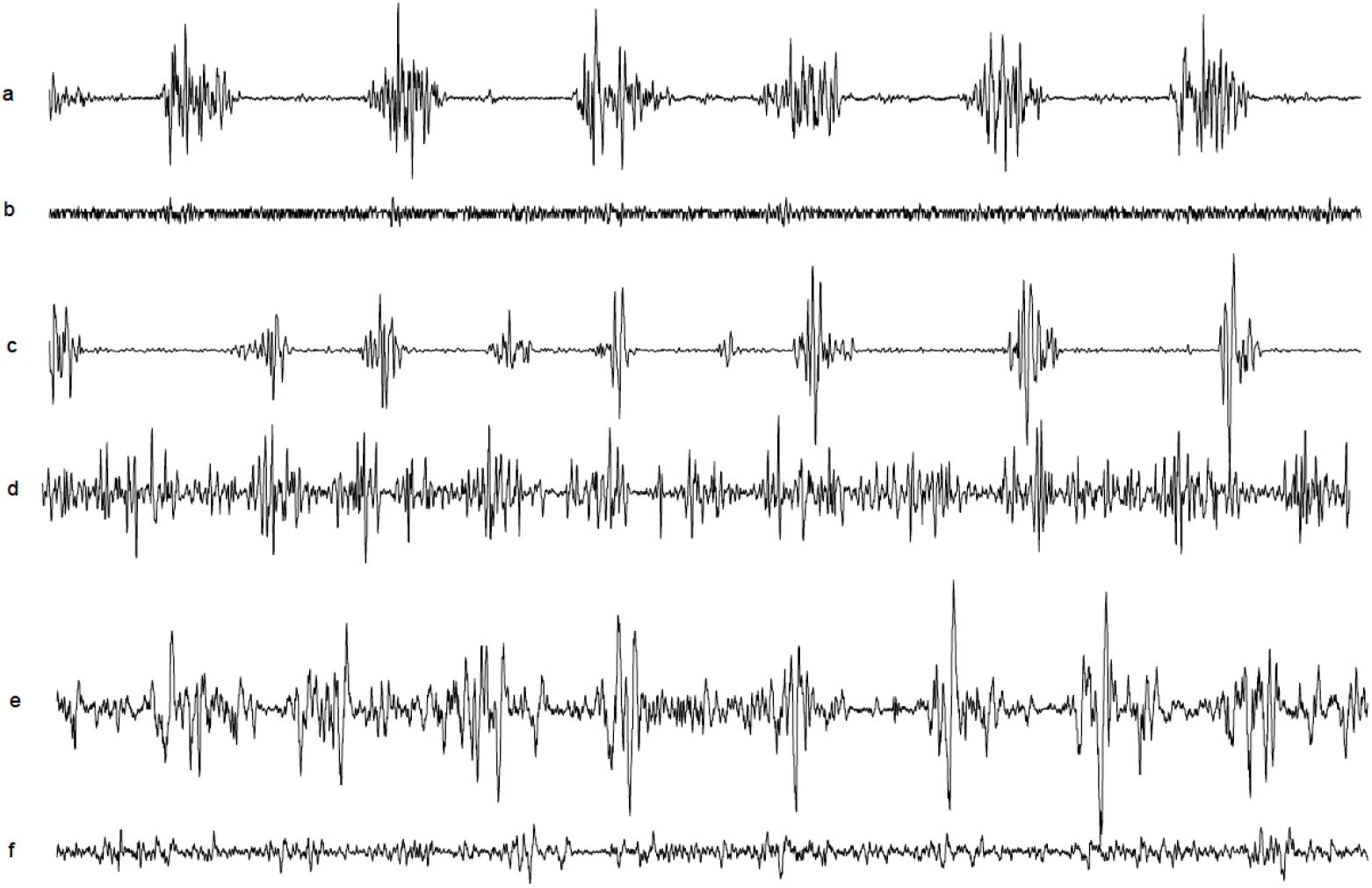
Representative surface electromyography findings in pseudo-orthostatic tremor. Representative surface electromyography findings in pseudo-orthostatic tremor. One-second surface electromyography (sEMG) excerpts from three patients with pseudo-orthostatic tremor (psOT). (a, b) Primary isolated psOT recorded from the right tibialis anterior (TA), showing 6.5-Hz tremor activity (a) that resolves with weight transfer (b). (c, d) Highly asymmetrical psOT, more prominent in the right rectus femoris (c) than in the left rectus femoris (d), presenting as an initial manifestation of Parkinson‚s disease (PD). (e, f) Functional psOT at 8 Hz (e), which resolves during a mental distraction task (f).

### Ancillary testing

Brain structural neuroimaging was performed in 25 patients (brain CT in 10, brain MRI in 16 and both in 1 patient). The results showed normal results in 15 of them and abnormalities, none of them deemed to be causative of psOT, in 10: Small vessel leukoencephalopathy was found in 9 patients and enlarged ventricles, in the absence of other features of hydrocephalus, in 1.

14 patients underwent a DaTSCAN SPECT, 2 of which, showed bilateral nigrostriatal denervation. 1 patient underwent a brain FDG-PET scan that showed hypometabolism in both parietal and temporal lobes as well as in the right precuneus, right occipital lobe and basal ganglia.

NCS/EMG were performed in 13 patients, 9 of which showed abnormal results: the findings in 4 patients, all in the organic psOT group, involved the lower limbs (diffuse axonal polyneuropathy in 3, chronic L4- L5-S1 denervation in 1). The remainder showed focal neuropathies in the median and/or ulnar nerves, that were judged as unrelated findings.

### Final clinical diagnoses

psOT was classified as functional in 6 (17%) and organic in 29 patients (83%) in our records, following the results of sEMG.

Nearly two thirds (65.5%) of the organic psOT could be classified as isolated tremor syndromes while only 9 (31%) were combined with other neurological or extraneurological manifestations that ranged from those clearly related to psOT, as is the case of parkinsonism, and those likely unrelated, such as hemifacial spasm. The clinical syndromes of organic psOT are illustrated in Figure 2. All functional psOT were classified as functional tremors.

**Figure 2.**
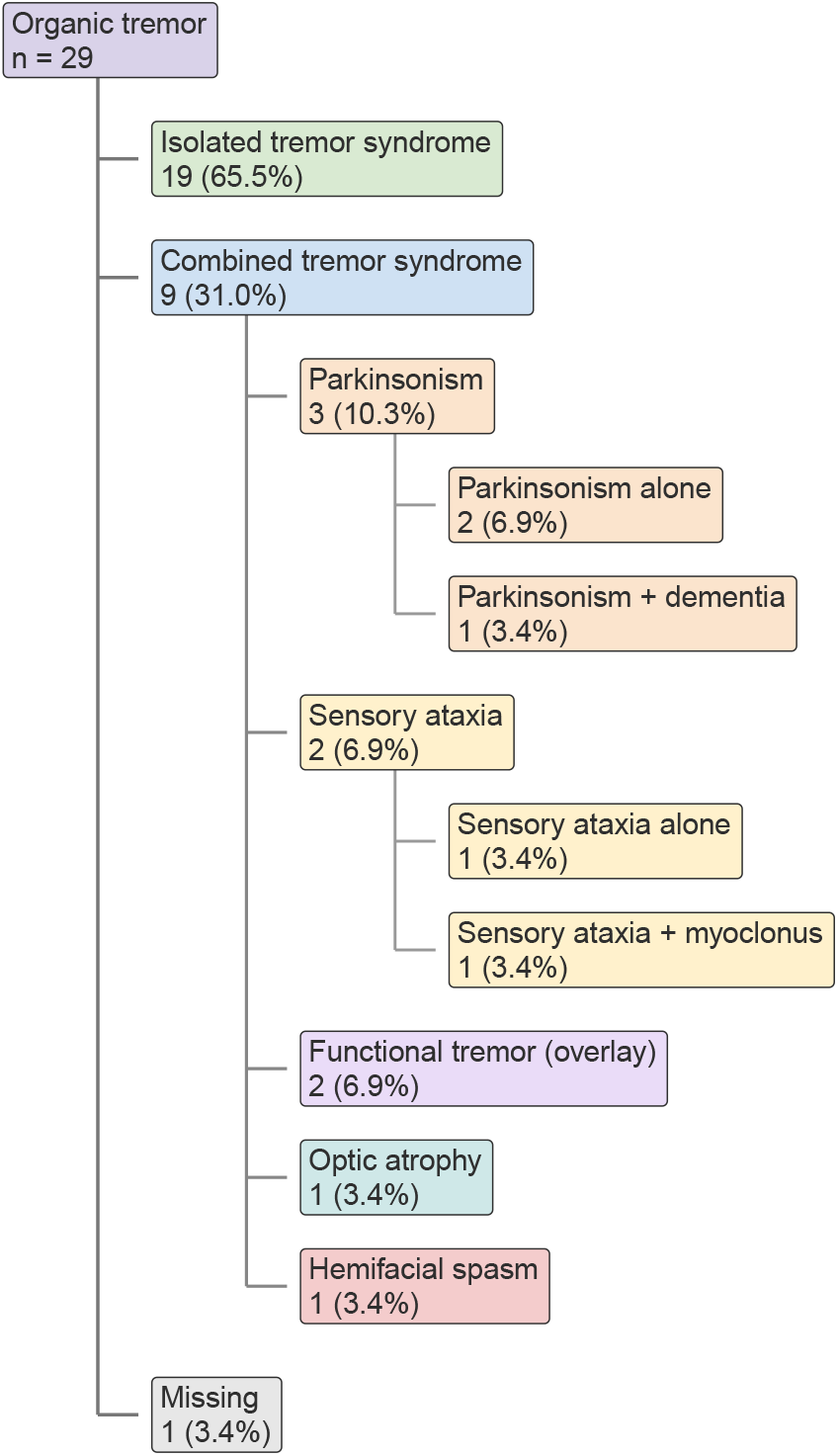
Hierarchical distribution of clinical syndromes in patients with organic tremor. Hierarchical distribution of clinical syndromes in the organic group. Numbers are presented as n (%), with percentages calculated using the total organic group (n = 29) as the denominator. Subcategories of combined tremor syndromes represent associated clinical features.

At the last follow-up, 21 of 29 patients with organic psOT (72.4%) fulfilled the diagnostic criteria for ET, independently of their standing-induced lower-limb tremor. An underlying neurological disorder to which psOT was considered attributable was identified in four patients (13.8% of organic cases): PD in two, Lewy body dementia (LBD) in one, and Leber hereditary optic neuropathy (LHON) in one. In the latter, genetic testing identified the mitochondrial MT-ND5 variant m.13042G>A (p.Ala236Thr), with 5% heteroplasmy. Two patients were classified as *primary isolated psOT*. In the remaining two patients, the etiology of psOT remained undetermined. One patient presented with significant unsteadiness associated with polyneuropathy and lumbar spinal stenosis, whereas the other had mixed-mechanism dementia (degenerative, vascular, and alcohol-related) and mild sensory axonal polyneuropathy. In neither case could the associated neurological conditions be confidently established as the cause of psOT.

Relevant comorbidities at last follow-up included chronic disabling dizziness in 1 organic patient, lumbar spinal stenosis in 2 organic patients, drug-induced exacerbation in 2 organic patients, complex FND in 2 functional patients and polyneuropathy in 3 and OM in 1 organic patients, respectively.

### Outcomes

Patient reported outcomes in the form of PGI-C were available for 25 patients, 20 organic and 5 functional (Supplementary Figure 3). The tremor improved in 4 (80%) of the functional and 11 (55%) of the organic patients but no statistically significant differences were found (p=0.615).

Among patients reporting improvement, ongoing pharmacological treatment included monotherapy with primidone (n=5), propranolol (n=3), clonazepam (n=1), or gabapentin (n=1), as well as combination therapy with clonazepam, gabapentin, and propranolol (n=1), or clonazepam, levodopa, opicapone, and rotigotine (n=1). Notably, only one patient in the functional psOT group had received pharmacological treatment for tremor (gabapentin), which was still being taken at the last follow-up.

The pharmacological treatments attempted and patient-reported outcomes are detailed in Supplementary table 3.

## Discussion

This retrospective cohort of patients with psOT is, to the best of our knowledge, the largest reported to date and the first to propose that psOT may be functional in a minority of patients and associated with ET in most.

In our series, 60% of all patients (72.4% of those with organic psOT) fulfilled the diagnostic criteria for essential tremor (ET), independently of their standing-induced lower-limb tremor, [6] at their last follow-up. This finding contrasts with previous reports emphasizing the association of psOT with other neurological disorders [15] and provides an opportunity to revisit the early hypothesis that slower forms of standing-induced lower-limb tremor may occur within the clinical spectrum of ET [4], [5] suggesting a possible overlap in the clinical phenomenology of POT and ET. Importantly, ET represents a clinical tremor syndrome (Axis I) rather than an etiological diagnosis (Axis II), and its coexistence with psOT should be interpreted within this framework. Interestingly, a family history of tremor was frequently reported among organic psOT patients with available information (15/19, 78.9%), although substantial missing data limit the interpretation of this finding. While familial aggregation is compatible with ET, family history is not a diagnostic criterion and cannot independently establish the relationship between ET and psOT.

The attenuation of tremor during weight transfer in 26 of 29 patients with organic psOT (90%) supports its weight-bearing nature. Furthermore, although balance and gait disturbances in ET have progressively gained attention [23], [24] the potential contribution of standing lower-limb tremor to them remains poorly understood. Interestingly, such balance disturbances were described relatively early [25] but were historically considered uncommon or attributed to aging rather than ET. Whether psOT contributes directly to unsteadiness or is a consequence of it remains uncertain and both possibilities have been considered in other orthostatic hyperkinesias [26], [27].

Our study also supports the fact that isolated organic psOT in the absence of other abnormalities, such as upper limb tremors, albeit infrequently, may indeed occur. On the other hand, the associations in secondary organic psOT largely align with those already reported in the literature, heralding the diagnosis of a potential alfa-synucleinopathy in 3 cases (PD in 2, LBD in 1 case) [13] and diagnosis of a mitochondrial disease in one. Whereas the association of LHON and tremor with or without other neurological features, termed LHON-plus, is relatively common [28] psOT had not been described. However, psOT has been previously linked to POLG, another mitochondrial disorder [29].

In our series, six patients (17%) were diagnosed with functional psOT on the basis of positive signs of internal inconsistency, supporting the recognition of functional psOT as a novel clinical category. Notably, neither of the two functional psOT patients with available weight-transfer data exhibited tremor attenuation during weight unloading. Furthermore, paradoxical worsening of tremor during weight unloading may represent an incongruent feature that should prompt clinicians to investigate other positive signs of functional tremor. Other findings, such as the apparent preferential involvement of thigh muscles in functional psOT, warrant further investigation, as their potential diagnostic significance remains uncertain. Although functional psOT has not, to our knowledge, been previously described, its identification is not entirely unexpected given the increasing recognition of the diverse clinical presentations of FND. Nevertheless, clinically recognizable signs suggestive of FND were documented in only three of the six patients (50%) with functional psOT, whereas four patients (13.8%) with organic psOT exhibited signs initially interpreted as functional but not subsequently confirmed by sEMG. These data outline the fact that FND presentations are protean and can mimic many different phenotypes [30], thus reinforcing the crucial need for a routine sEMG study.

Despite its strengths, our study does not come without limitations. First, its retrospective design is sensitive to missing data. sEMG data, while valuable, have been extracted from the records and not from the neurophysiological raw data which has led to a lack of data on relevant features such as coherence or burst duration. In addition, patient reported outcomes were collected retrospectively and are subject to bias while treatment strategies likely reflect trial and error strategies which limits our ability to draw conclusions. Last, some issues remain unsolved such as the relationship between contributors to unsteadiness (e.g. polyneuropathy) and tremor generation or the cases with functional overlay, that might reflect true overlapping between FND and organic mechanisms or our inability to demonstrate internal inconsistencies in these cases.

Our research presents some clinical implications: 1) The strong association between organic psOT and ET, representing 60% of the cohort, challenges the prevailing view that psOT is almost always part of a combined tremor syndrome and attributable to a known neurological disorder 2) The 90% rate of tremor attenuation during weight transfer in organic cases supports a weight-bearing mechanism, possibly questioning the current nosology. 3) Defining functional psOT (17% of cases) as a novel category characterized by sEMG signs of internal inconsistency underscores the diagnostic necessity of routine electrophysiological studies. 4) Recognizing organic psOT as a potential early clinical manifestation of alpha-synucleinopathy or mitochondrial pathology is critical for proactive neurological screening.

Large-scale, ideally multicenter prospective studies are required to validate these retrospective findings and capture raw neurophysiological parameters, such as coherence and burst duration. Also, the bidirectional relationship between lower-limb tremor and postural instability must be systematically investigated to determine if tremor drives unsteadiness or acts as a compensatory response (or both). Finally, future research must clarify the mechanisms of functional overlay and the modulating role of comorbid peripheral or structural neuropathies in lower-limb tremor generation.

## Conclusions

Our retrospective study suggests that psOT is most often associated with ET, while an identifiable neurological cause was found in a relatively small proportion of patients. In these patients psOT often behaves like a true weight-bearing hyperkinesia, potentially contradicting its current nosology. Rare cases of isolated psOT without an identifiable underlying etiology were also observed, provisionally designated as *primary isolated psOT*. Additionally, we have shown that psOT can be a presentation of FND and that its recognition relies on demonstrable internal inconsistency in sEMG studies. There is a need for larger scale prospective studies to better delineate the clinical and neurophysiological spectrum of psOT phenomena.

## Statements and declarations

### Data availability

The data supporting the findings of this study are not publicly available due to patient confidentiality and privacy considerations. Anonymized data may be made available from the corresponding author upon reasonable request.

## CRediT authorship contribution statement

- Agustín Querejeta-Coma: Conceptualization, Methodology, Investigation, Data curation, Formal analysis, Project administration, Supervision, Visualization, Writing – original draft, Writing – review & editing.
- Luis Yupanqui-Guerra: Investigation, Data curation, Writing – review & editing.
- Laura Santos-Sánchez-de-las-Matas: Investigation, Data curation, Writing – review & editing.
- Adriana Gómez-Domínguez: Investigation, Data curation.
- Héctor Canfield-Medina: Investigation, Data curation.
- Roberto Cano-de-la-Cuerda: Conceptualization, Methodology, Writing – review & editing.
- Carlos Ordás-Bandera: Conceptualization, Methodology, Investigation, Data curation, Project administration, Supervision, Visualization, Writing – review & editing.

## Declaration of generative AI and AI-assisted technologies in the manuscript preparation process

During the preparation of this manuscript, the authors used ChatGPT (OpenAI) to assist with language editing, clarity, and manuscript organization. The authors subsequently reviewed and edited the content and take full responsibility for the final manuscript.

