## Supplementary material for "Functional and Organic Pseudo-Orthostatic Tremor: Clinical Features, Electrophysiology, and Treatment Outcomes in a Retrospective Cohort"

**Supplementary information**

Supplementary Figure 1 Prior use of tremor-inducing medications in functional and organic pseudo-orthostatic tremor

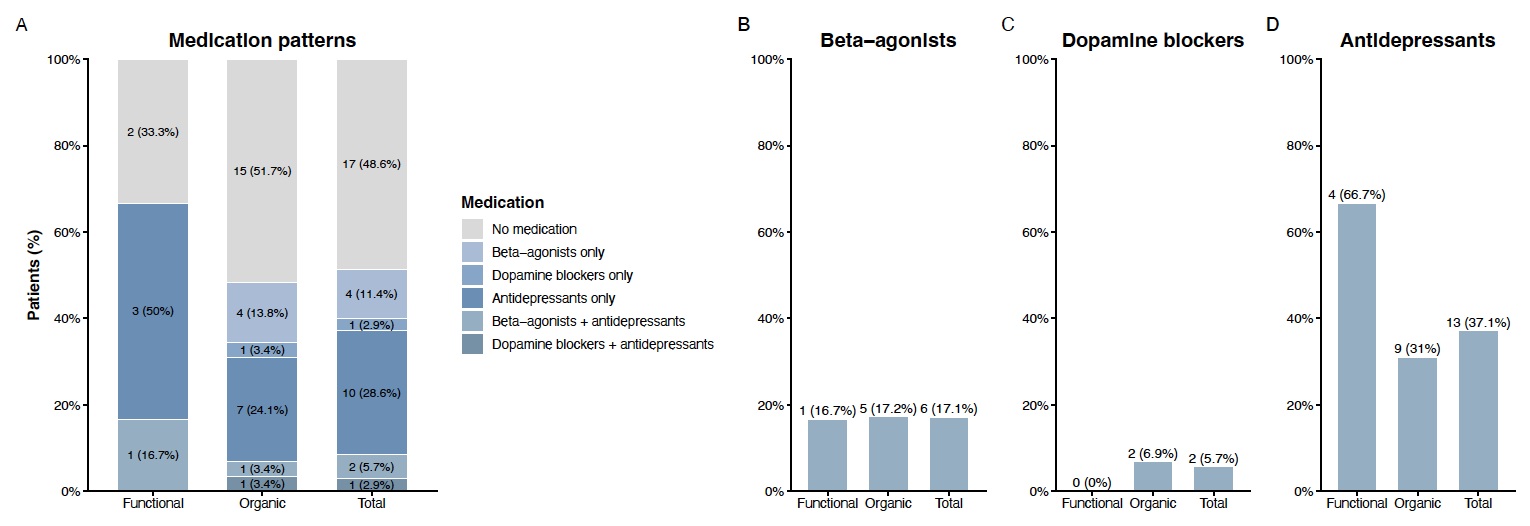

(A) Distribution of medication exposure patterns according to drug class. (B–D) Proportion of patients with previous exposure to beta-agonists, dopamine blockers, and antidepressants, respectively. Values are shown as n (%). Total refers to the entire series.

Supplementary Table 1: Clinical characteristics of organic and functional pseudo-orthostatic tremor

| Patients (n) | Organic pseudo-OT (29) | Functional pseudo-OT (6) | All (35) | *p-*value |
| --- | --- | --- | --- | --- |
| Symptoms |  |  |  |  |
| Lower limb tremor, n (%) | 23 (79) | 4 (67) | 27 (77) | 0.602 |
| Trunk tremor, n (%) | 0 | 3 (50) | 3 (8.6) | 0.003 |
| Unsteadiness, n (%) | 16 (55) | 4 (67) | 20 (57) | 0.68 |
| Lower limb weakness, n (%) | 3 (10) | 0 | 3 (8.6) | 1 |
| Falls, n (%) | 10 (35) | 2 (33) | 12 (34) | 1 |
| Fear of falling without falls, n (%) | 1 (3.4) | 0 | 1 (2.9) | 1 |
| Back pain, n (%) | 2 (6.9) | 0 | 2 (5.7) | 1 |
| Orthostatic intolerance, n (%) | 0 | 1 (17) | 1 (2.9) | 0.171 |
| Upper limb tremor, n (%) | 24 (83) | 2 (33) | 26 (74) | 0.027 |
| Head tremor, n (%) | 6 (21) | 0 | 6 (17) | 0.56 |
| Voice tremor, n (%) | 2 (7) | 0 | 2 (5.7) | 1 |
| Face tremor, n (%) | 2 (7) | 0 | 2 (5.7) | 1 |
| Examination findings |  |  |  |  |
| Lower limb tremor, n (%) | 26 (90) | 4 (67) | 30 (86) | 0.195 |
| Romberg’s sign, n (%) | 4 (14) | 0 | 4 (11) | 1 |
| Unsteady gait, n (%) | 5 (17) | 0 | 5 (14) | 0.561 |
| Parkinsonism, n (%) | 1 (3.4) | 0 | 1 (2.9) | 1 |
| Upper limb action tremor, n (%) | 22 (76) | 3 (50) | 25 (71) | 0.322 |
| Upper limb resting tremor, n (%) | 2 (7) | 0 | 2 (5.7) | 1 |
| Myoclonus, n (%) | 1 (3.4) | 0 | 1 (2.9) | 1 |
| Head tremor, n (%) | 3 (10) | 0 | 3 (8.6) | 1 |
| Voice tremor, n (%) | 1 (3) | 0 | 1 (2.9) | 1 |
| Lower limb weakness, n (%) | 1 (3) | 0 | 1 (2.9) | 1 |
| Positive signs of FND, n (%) | 4 (14) | 3 (50) | 7 (20) | 0.079 |

FND: Functional neurological disorder; OT: orthostatic tremor

Supplementary Figure 2: Distribution of the tremor across the lower limbs in both organic and functional pseudo-orthostatic tremor

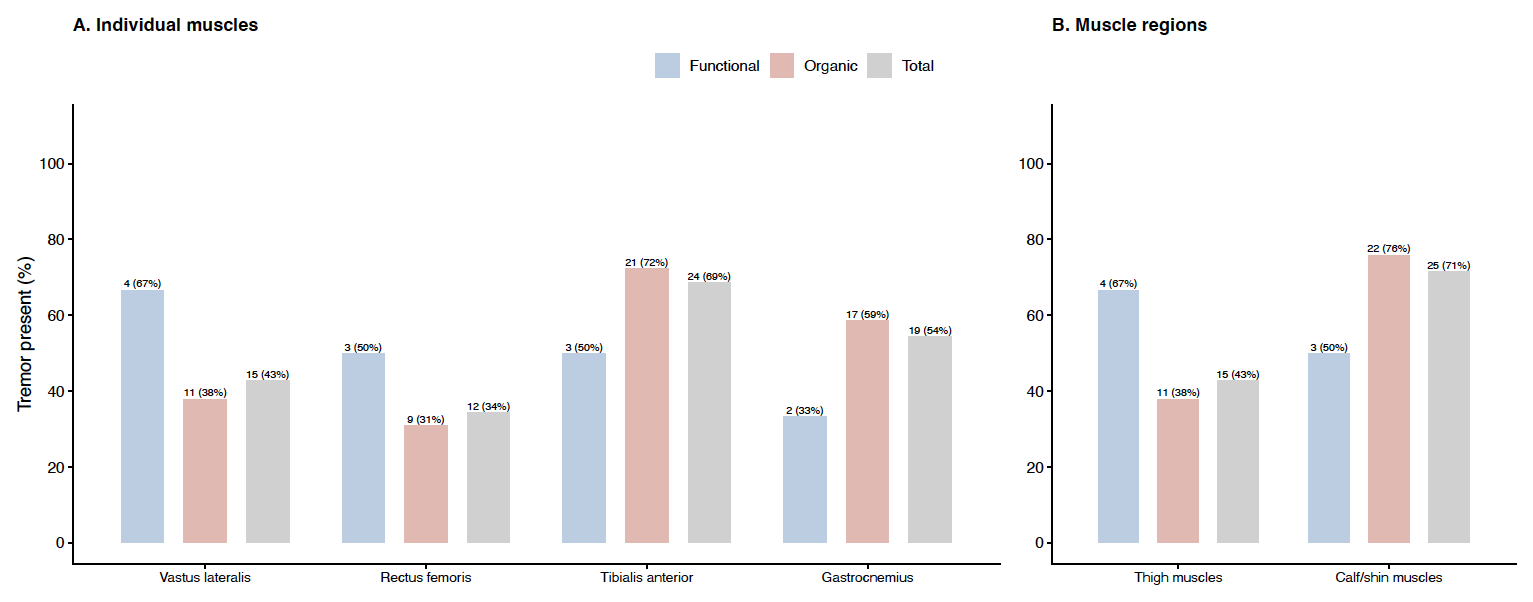
Bars represent the number and percentage of patients with tremor recorded in each muscle or muscle region. Thigh involvement was defined as tremor in the vastus lateralis and/or rectus femoris, and calf/shin involvement as tremor in the tibialis anterior and/or gastrocnemius. “Total” refers to the entire series.

Supplementary Figure 3. Patient Global Impression of Change by tremor group

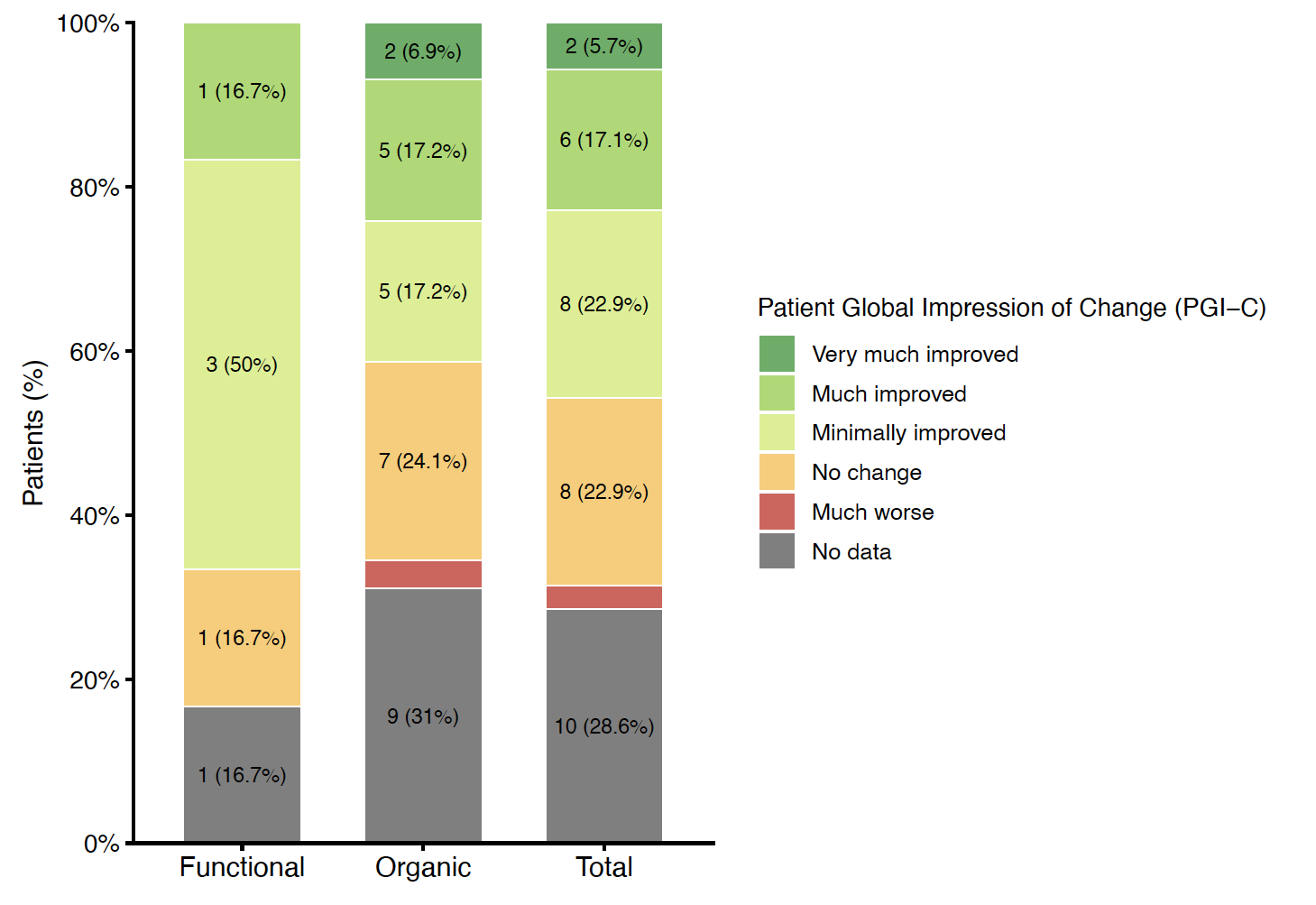

Distribution of Patient Global Impression of Change (PGI-C) ratings in patients with functional and organic tremor and in the overall cohort. Bars represent the percentage of patients in each PGI-C category; values within segments are shown as n (%). Patients without an available PGI-C assessment are included as “No data.”

| **Drug class** | **Specific agent** | **Functional, n** | **Organic, n** |
| --- | --- | --- | --- |
| Beta-agonists | Budesonide/formoterol | 0 | 2 |
|  | Salbutamol | 0 | 2 |
|  | Formoterol | 1 | 0 |
|  | Indacaterol | 0 | 1 |
|  | Terbutaline | 0 | 1 |
| Dopamine blockers | Clebopride | 0 | 1 |
|  | Levosulpiride | 0 | 1 |
| Antidepressants | Duloxetine | 2 | 2 |
|  | Fluoxetine | 0 | 3 |
|  | Mirtazapine | 1 | 2 |
|  | Escitalopram | 0 | 2 |
|  | Citalopram | 1 | 0 |
|  | Desvenlafaxine | 0 | 1 |
|  | Sertraline | 0 | 1 |
|  | Venlafaxine | 1 | 0 |
| Other | Prednisone | 1 | 0 |

Supplementary table 2. Previous exposure to potentially tremor-inducing medications by study group

Values are presented as number of patients (n). Individual patients may have been exposed to more than one medication and may therefore contribute to more than one row. Budesonide/formoterol was classified under beta-agonists.

Supplementary Table 3. Pharmacological treatments attempted and patient-reported outcomes in functional and organic pseudo-orthostatic tremor

| Medication | Organic (n=29) | Functional (n=6) | Total (n=35) |
| --- | --- | --- | --- |
| Bisoprolol | 0/1 (0.0%) | 0/0 (—) | 0/1 (0.0%) |
| Clonazepam | 3/8 (37.5%) | 0/0 (—) | 3/8 (37.5%) |
| Gabapentin | 2/7 (28.6%) | 1/1 (100.0%) | 3/8 (37.5%) |
| Levetiracetam | 0/1 (0.0%) | 0/0 (—) | 0/1 (0.0%) |
| Levodopa | 1/1 (100.0%) | 0/0 (—) | 1/1 (100.0%) |
| Opicapone | 1/1 (100.0%) | 0/0 (—) | 1/1 (100.0%) |
| Perampanel | 0/4 (0.0%) | 0/0 (—) | 0/4 (0.0%) |
| Pregabalin | 0/1 (0.0%) | 0/0 (—) | 0/1 (0.0%) |
| Primidone | 5/10 (50.0%) | 0/0 (—) | 5/10 (50.0%) |
| Propranolol | 4/11 (36.4%) | 0/0 (—) | 4/11 (36.4%) |
| Rotigotine | 1/1 (100.0%) | 0/0 (—) | 1/1 (100.0%) |
| Zonisamide | 0/4 (0.0%) | 0/0 (—) | 0/4 (0.0%) |

Data are presented as effective/attempted treatments (%). A treatment was considered attempted if it was currently being used or had previously been discontinued. Treatment was considered effective when it was currently being used and the patient reported improvement on the Patient Global Impression of Change (PGI-C; minimally improved, much improved, or very much improved). Discontinued treatments and current treatments without reported improvement were considered ineffective. A dash indicates that the treatment was not attempted by any patient in that group.
